# COVID-19 containment policies and hyperglycemia in pregnancy: correlation with the Stringency Index in a nationwide Belgian cohort

**DOI:** 10.64898/2026.06.17.26355901

**Authors:** Elena Costa, An Vercoutere, Sophie Alexander, Michel Boulvain, Sara Derisbourg, Clotilde Lamy, Anne Delbaere, Judith Racapé

## Abstract

**Introduction:** During the COVID-19 pandemic, gestational diabetes mellitus (GDM) prevalence showed variable changes across regions, with most reporting increases and others decreases; anxiety and lifestyle modifications linked to pandemic-related restrictions have been proposed as potential contributors in case of increase, while changes to less stringent screening strategies in case of decrease. The correlation of the GDM variation according to the intensity of pandemic restrictions has not been described to date. Our study aimed to examine the correlation between hyperglycemia in pregnancy (HIP) prevalence and pandemic-related restrictions measured by the Stringency Index (SI) and compare neonatal weight percentiles between the pre pandemic and the pandemic period.

**Materials and Methods:** We included all singleton live births in Belgium in 2019 and 2020 from Belgian birth registry data (n=229,170). We compared monthly proportions of HIP prevalence and Small for gestational age (SGA) and Large for gestional age (LGA) newborns in 2019 and 2020. Crude and adjusted odds ratios (ORs, aORs) for socio-economic status, maternal age, parity and BMI were estimated with logistic and multinomial regression. The Spearman correlation coefficient was used to assess the correlation between the monthly average SI and the monthly aORs of HIP.

**Results:** For deliveries from January to June 2020, no significant differences in HIP prevalence were observed compared with 2019. For births occurred July to December 2020, there was a significant increase in HIP, with peaks in July (GDM screening in April) (aOR 1.41, 1.26–1.58) and November (GDM screening in August) (aOR 1.33, 95% CI 1.18–1.49). The Spearman correlation coefficient between the SI and HIP aORs was 0.82 (p = 0.01).There was no significant change in neonatal weight percentiles.

**Conclusion:** During the pandemic, we observed an increase in the prevalence of HIP and its increase during the was strongly correlated with the corresponding SI. There was no measurable impact on the frequency of LGA or SGA newborns. These results may be useful to anticipate unintended consequences of future public health emergencies requiring similar containment measures.

## Introduction

During the COVID-19 pandemic, several studies reported changes in the prevalence of gestational diabetes mellitus (GDM), with most describing an increase in prevalence and a smaller number reporting decreases, mostly related to modifications in screening strategies.(1–7).

Although the mechanisms underlying these changes remain incompletely understood, anxiety and lifestyle modifications linked to pandemic-related restrictions have been proposed as potential contributors to the increase of GDM prevalence(2,4,8).

Containment measures implemented during the pandemic may influence glucose metabolism through several pathways. Restrictions on movement and social activities may reduce opportunities for physical activity(9,10), alter dietary habits(10), and contribute to excessive gestational weight gain(1). In addition, uncertainty, social isolation, and concerns regarding health and access to care may increase psychological stress and anxiety(11). These factors have all been hypothesized to contribute to impaired glucose regulation during pregnancy and a higher risk of GDM and more broadly hyperglycaemia in pregnancy (HIP).

HIP is a well-established risk factor for excessive fetal growth and large-for-gestational-age (LGA) newborns (12,13). Therefore, if pandemic-related restrictions were associated with an increase in HIP prevalence, one might expect a corresponding increase in the proportion of LGA newborns at the population level. However, previous studies have generally reported little or no change in birthweight-related outcomes during the pandemic(3,14–16).

The intensity of governmental containment measures during the COVID-19 pandemic has been quantified using the Stringency Index (SI), that captures the severity of restrictions such as school closures, workplace closures, gathering limitations, stay-at-home requirements, and travel restrictions. (17), The SI offers a standardized way to compare the intensity of public health restrictions over time and across settings.

Although several studies have examined changes in gestational diabetes prevalence during the pandemic, most have focused on comparisons between pre-pandemic and pandemic periods. To our knowledge, no study has directly investigated whether the intensity of governmental containment measures, quantified through the SI, was associated with temporal variations in HIP prevalence.

Beyond the context of COVID-19, understanding how pregnancy-related conditions respond to periods of restricted mobility and social disruption may be relevant for future public health emergencies. Future pandemics, natural disasters, environmental crises, or other situations requiring movement restrictions could produce similar indirect effects on maternal health.

Identifying associations between restriction intensity and pregnancy outcomes may therefore help anticipate risks and guide preventive strategies during future emergencies.

The primary objective of our study was to evaluate the prevalence of HIP in 2020 compared with 2019 and the correlation between the SI and HIP prevalence variation. The secondary objectives were to evaluate the distribution of small for gestational age (SGA) and large for gestational age (LGA) in 2020 compared with 2019.

## Materials and methods

### Study population and data

We used data from birth certificates collected from the Belgian civil registration system. We included all singleton babies born in Belgium between the 1^st^ of January 2019 and the 31^st^ December 2020. Medical data included in the registration system were recorded at the hospital by midwives and gynecologists. Sociodemographic data were recorded by the civil registration service within a fortnight of birth, as reported by the parent(s). In Belgium, two perinatal epidemiological centres, SPE (Studiecentrum voor Perinatale Epidemiologie-Study Centre for Perinatal Epidemiology) for the Dutch-speaking region and CEpiP (Centre d’Épidémiologie Périnatale - Center for Perinatal Epidemiology) for the French-speaking region, are in charge of data quality and completeness, as catalogued in birth and death certificates.

Registry data for this study were first accessed on June 20^th^ 2024. Data were received already pseudonymised.

### Outcomes and variables definitions

#### Outcomes

HIP includes both gestational and preexisting diabetes, although most cases are likely gestational diabetes due to the low prevalence of type 1 and type 2 diabetes in European women of reproductive age (0.3% and 0.2%, respectively)(18). In the birth certificates, HIP was reported at the time of delivery. We considered though that gestational diabetes was diagnosed in the month in which gestational diabetes screening was most likely performed, according to guidelines in vigour at the time (19,20).

We calculated birthweight percentiles for newborns via AUDIPOG (Association des Utilisateurs de Dossiers Informatisés en Pédiatrie, Obstétrique et Gynécologie)(21) curves, categorising them as below the 10th percentile (SGA), between the 10th and 90th percentiles (Adequate for Gestational Age–AGA), and above the 90th percentile (LGA).

#### Variable definitions

**The SI** (17) is a measure of the strictness of governmental containment policies during the COVID-19 pandemic. It is a composite measure based on nine indicators: school closures, workplace closures, cancellation of public events, restrictions on public gatherings, closures of public transport, stay-at-home requirements, public information campaigns, restrictions on internal movements, and international travel controls. The index ranges from 0 to 100, with 100 being the strictest restrictions and 0 being the absence of restrictions. If policies varied at the subnational level, the index reflected the level of the strictest subregion.

**Maternal nationality of origin** was defined as the nationality of the mother at the time of her birth. We grouped nationalities into the seven most represented categories: Belgium, European Union 15, European Union 27 excluding European Union 15, Eastern Europe, North Africa, Sub-Saharan Africa, Turkey, and "Other."

**Sociodemographic variables** (SDVs) included maternal age, parity, maternal education level, single women, and the number of incomes in the household. Maternal age was categorised into three groups: under 20 years, 20-35 years, and 35 years and over. Parity was grouped into three categories: 1, 2 to 3, and 4 or more births. Maternal education was divided into three categories: higher (university or higher education), secondary (completed secondary school), and primary (completed primary school or less) education level. Single women were defined as those not living with a partner. Number of incomes in the household was determined by combining paternal and maternal occupational statuses, resulting in categories of two, one, or no income. For single women, the number of incomes could vary between zero and one.

Maternal body mass index (BMI) was calculated from weight and height recorded at the first antenatal consultation. The BMI categories were underweight (<18.5 kg/m²), normal weight (18.5–24.99 kg/m²), overweight (25–30 kg/m²), and obese (>30 kg/m²).

Hypertension (HT) encompassed all diagnoses related to maternal hypertension (preexisting HT, pregnancy-induced HT, preeclampsia).

### Statistical methods

Descriptive statistics for baseline characteristics and for pregnancy and newborn outcomes (hypertension, HIP, and newborn weight percentiles), were presented with counts and percentages in 2019 and 2020. We compared the percentage by using chi² Pearson test We compared HIP prevalence in 2020 and 2019 month-by-month. We reported crude odds ratios (ORs) and 95% confidence intervals (CIs) via univariate logistic regression and adjusted the ORs for baseline characteristics (maternal nationality of origin, number of incomes within the family, being single, maternal education, maternal age, parity, maternal BMI) via multivariate logistic regression.

To analyse the correlation between adjusted odds ratios (aORs) for HIP in 2020 versus 2019 and the SI, values were assigned based on the month of screening. Given that screening occurred between 24 and 28 weeks’ gestation (11,12), the timing was approximated to 26 weeks’ gestation for analytical purposes. We then used Spearman’s correlation coefficient to analyse the relationship between the aORs of HIP and the average SI for each month.

To investigate the impact of the pandemic on birthweight percentiles, we compared the proportions of SGA, AGA and LGA newborns between 2020 and 2019. We calculated crude odds ratios (ORs) and 95% CIs via univariate multinomial logistic regression month-by-month. We adjusted the ORs for baseline and pregnancy characteristics (maternal BMI, parity, single, household incomes, maternal nationality of origin, maternal age, presence of hypertension) via multivariate multinomial logistic regression.

We present the odds ratios and 95%CI derived from the logistic regression models and the p-values for the Wald *χ*^2^ test. The significance level was set at α = 0.05, and all analyses were performed using Stata SE15 software Plots were produced via Python. Microsoft Co-Pilotwas used for helping with Python coding for plot generation and for increasing the readability of the text.

### Ethics statement

This study and the use of related data were approved by the Institutional Review Board (IRB) of the Belgian Statistical Office (Statbel): reference number 2022/052.

Participant consent is the responsibility of Statbel in accordance with Belgian legislation(22).

## Results

### Characteristics of the population in 2019 and 2020

This study included all singleton deliveries that occurred in 2019 (n=116351) and 2020 (n=112819), totalling 229170 deliveries. The baseline characteristics of the population in 2019 and 2020 are presented in Table 1.

**Table 1.** Baseline characteristics of the studied population in 2019 and 2020.

|  | 2019 |  | 2020 |  | p value |
| --- | --- | --- | --- | --- | --- |
|  | n | % | n | % |  |
| **Maternal Age** |  |  |  |  |  |
| <25 years | 16329 | 14.05% | 15961 | 14.15% | 0.63 |
| 25-35 years | 81477 | 70.11% | 79090 | 70.13% |  |
| >35 years | 18401 | 15.83% | 17723 | 15.72% |  |
| **Nationality at birth** |  |  |  |  |  |
| Belgium | 56361 | 60.86% | 58875 | 61.96% | <0.001 |
| EU15 | 7350 | 7.94% | 7599 | 8% |  |
| EU27 without EU15 | 4793 | 5.18% | 4733 | 4.98% |  |
| Eastern Europe non-EU27 | 2563 | 2.77% | 2637 | 2.78% |  |
| Turkey | 2199 | 2.37% | 2116 | 2.23% |  |
| Northern Africa | 8603 | 9.29% | 8331 | 8.77% |  |
| Sub-Saharan Africa | 5566 | 6.01% | 5478 | 5.76% |  |
| Other | 5177 | 5.59% | 5255 | 5.53% |  |
| **Maternal education** |  |  |  |  |  |
| Primary or less | 5113 | 5.33% | 3587 | 4.18% | <0.001 |
| Secondary | 46221 | 48.19% | 41114 | 47.94% |  |
| Higher education | 44582 | 46.48% | 41067 | 47.88% |  |
| **Single Status** |  |  |  |  |  |
| In a couple | 97861 | 86.01% | 95725 | 86.18% | 0.26 |
| Not in a couple | 15912 | 13.99% | 15352 | 13.82% |  |
| **Number of incomes in the family household** |  |  |  |  |  |
| 0 | 13679 | 13.49% | 10846 | 11.68% | <0.001 |
| 1 | 24753 | 24.42% | 22868 | 24.62% |  |
| 2 | 62948 | 62.09% | 59179 | 63.71% |  |
| **Parity** |  |  |  |  |  |
| Primiparous | 49845 | 43.39% | 49934 | 44.57% | <0.001 |
| 1 or two births | 55363 | 48.82% | 52960 | 47.28% |  |
| 3 births or more | 9664 | 8.41% | 9129 | 8.15% |  |
| **BMI** |  |  |  |  |  |
| Underweight | 5426 | 4.94% | 5062 | 4.76% | <0.001 |
| Normal weight | 60420 | 54.99% | 57738 | 54.31% |  |
| Overweight | 28006 | 25.49% | 27350 | 25.73% |  |
| Obese | 16028 | 14.59% | 16160 | 15.2% |  |

Baseline characteristics were largely comparable between 2019 and 2020. Maternal age and couple status did not differ significantly between the two years. Statistically significant differences were observed for maternal nationality at birth, educational level, household income, parity, and BMI distribution; however, the absolute differences were small. In particular, the proportion of women with higher education, living in households with two incomes, and classified as obese increased slightly in 2020 compared with 2019.

With respect to pregnancy characteristics and newborn outcomes (Table 2), the distribution of birthweight percentiles was similar in 2019 and 2020, with no significant differences in the proportions of SGA, AGA or LGA newborns. Hypertension prevalence was slightly lower in 2020 than in 2019 (4.48% vs 4.69%, p=0.02). In contrast, the prevalence of HIP increased from 8.89% in 2019 to 10.26% in 2020 (p<0.001).

**Table 2.**
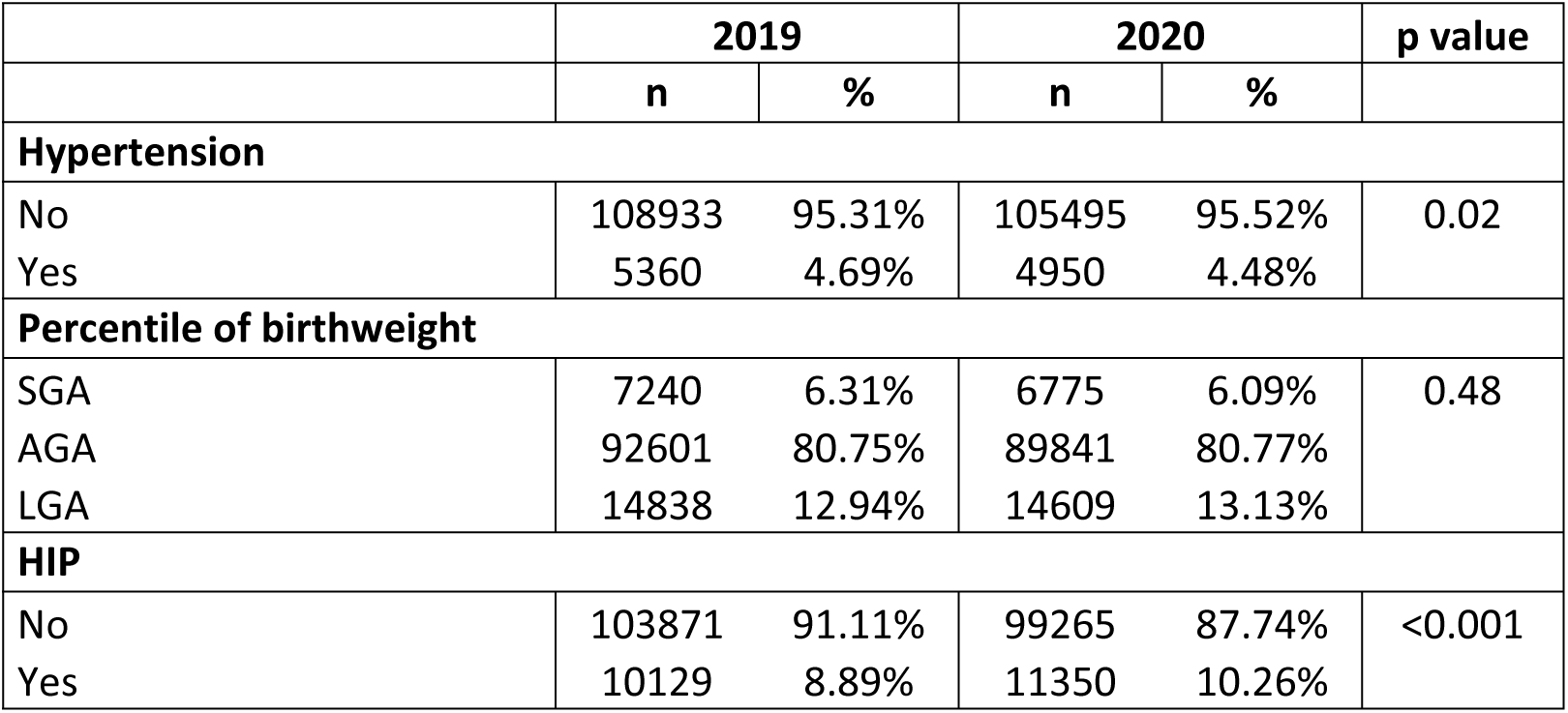
Pregnancy characteristics and newborn outcomes in 2019 and 2020.

### HIP by month of delivery in 2019 et 2020

When stratified by month of delivery, we observed that for patients who delivered from January to June, there were no statistically significant differences in the prevalence of HIP between 2020 and 2019. For patients who delivered from July to December, there was a significant increase in HIP in 2020 compared with the same months in 2019, with two peaks in July (aOR 1.41 95% CI 1.26–1.58) and November (1.33 95% CI 1.18–1.49) (Table 3).

**Table 3.** Proportion of pregnancies affected by HIP, Odds Ratios (CI95%) and adjusted Odd Ratio (CI95%) stratified by month if delivery in 2019 and 2020. * p<0.05 ° *Adjustment for maternal nationality of origin, household incomes, single status, maternal education, maternal age, parity, maternal BMI*

| Month of delivery | HIP 2020 |  | HIP 2019 |  | OR (95%CI) | aOR (95%CI)° |
| --- | --- | --- | --- | --- | --- | --- |
|  | n | % | n | % |  |  |
| January | 834 | 8,85 | 781 | 8,03 | **1.11(1-1.23)\*** | 1.05(0.93-1.19) |
| February | 763 | 8,99 | 693 | 8,12 | **1.11(1-1.24)\*** | 1.04(0.92-1.19) |
| March | 834 | 8,91 | 709 | 7,9 | **1.12(1-1.26)\*** | 1.06(0.93-1.21) |
| April | 773 | 8,35 | 789 | 8,44 | 0.98(0.89-1.1) | 0.97(0.85-1.11) |
| May | 803 | 8,66 | 788 | 8,13 | 1.07(0.96-1.18) | 1(0.88-1.14) |
| June | 894 | 9,5 | 794 | 8,34 | **1.15(1-1.27)\*** | 1.09(0.96-1.23) |
| July | 1188 | 11,75 | 921 | 8,83 | **1.37(1.25-1.5)\*** | **1.41(1.26-1.58)\*** |
| August | 1095 | 11,58 | 983 | 9,87 | **1.15(1.05-1.26)\*** | **1.18(1.06-1.32)\*** |
| September | 1069 | 11,33 | 960 | 9,94 | **1.15(1-1.26)\*** | **1.16(1.04-1.29)\*** |
| October | 1062 | 11,29 | 966 | 9,77 | **1.17(1.1-1.28)\*** | **1.13(1.01-1.26)\*** |
| November | 1033 | 12,14 | 865 | 9,47 | **1.32(1.2-1.45)\*** | **1.33(1.18-1.49)\*** |
| December | 1002 | 11,81 | 880 | 9,64 | **1.25(1.14-1.38)\*** | **1.22(1.09-1.36)\*** |

### Association of HIP with stringency index

The Spearman correlation coefficient between the average stringency index by month and the aOR of HIP in 2020, compared to 2019, was 0. 82 (p = 0.01) (Fig1).

**Figure 1.**
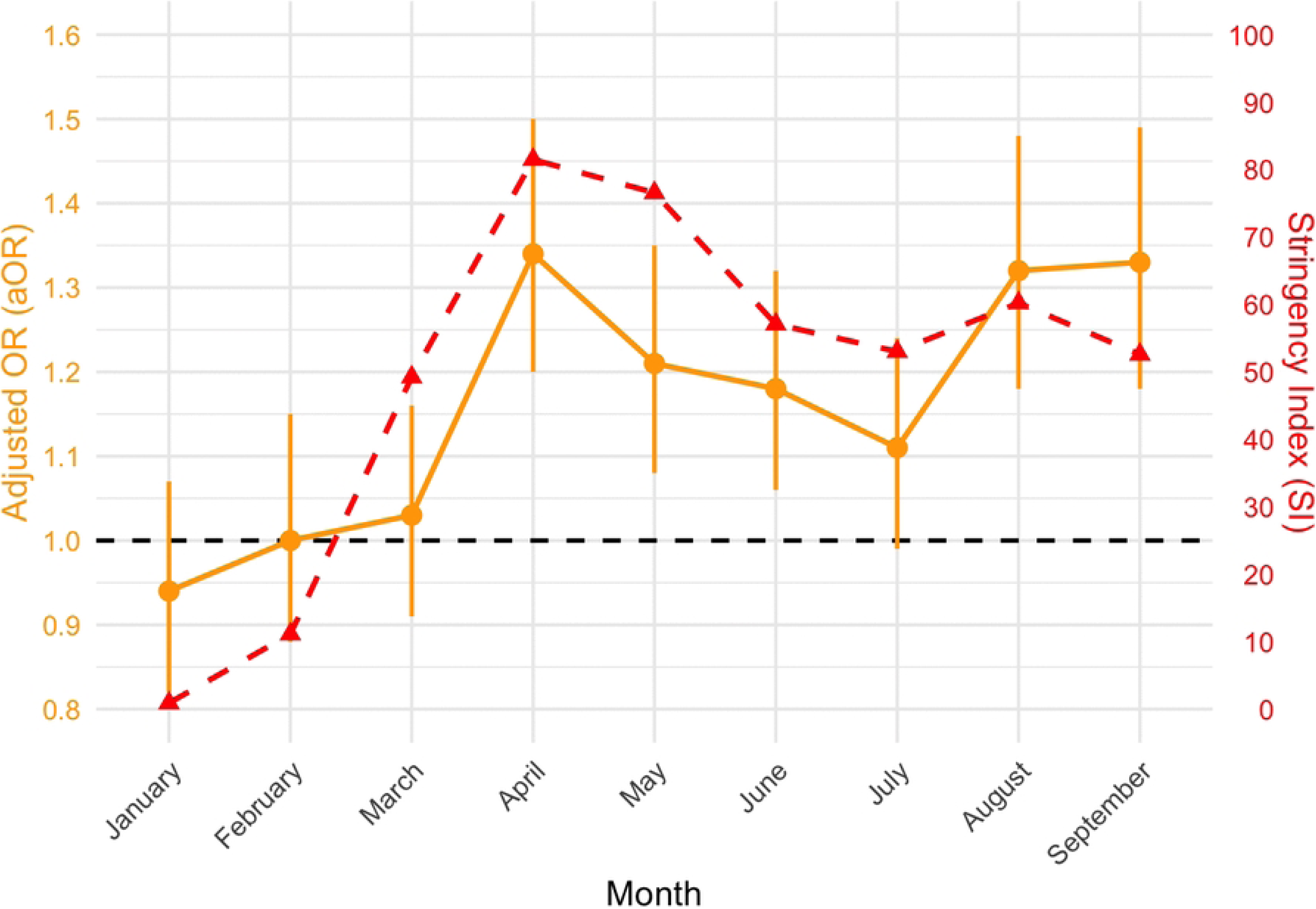
Plot of the adjusted odds ratios per month of screening of HIP and the average stringency index. *The aOR values (2020 compared to 2019) are reported for the month of screening approximated as 26 weeks of gestation. The x-axis represents the months of the year, while the primary y-axis on the left indicates the aOR values, and the secondary y-axis on the right indicates the average stringency index values*.

### Newborn weight percentile by month of delivery in 2019 et 2020

There was no statistically significant difference in the distribution of newborn weight percentile according to the month of delivery, with the exception of a reduction in the proportion of LGA newborns in the month of September 2020 compared with September 2019 (aOR 0.89 95% CI 0.8--0.99) (Table 4).

**Table 4.**
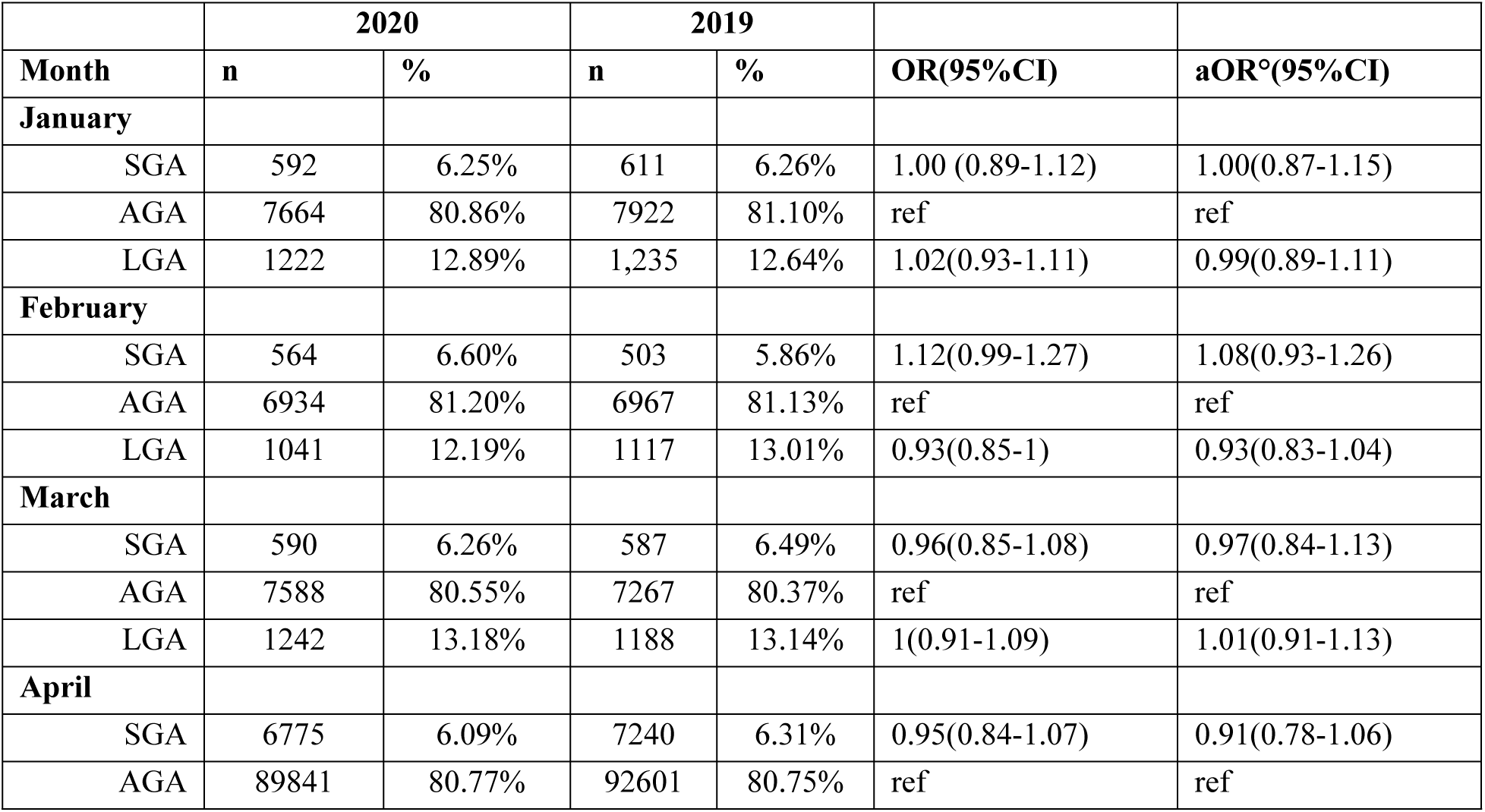
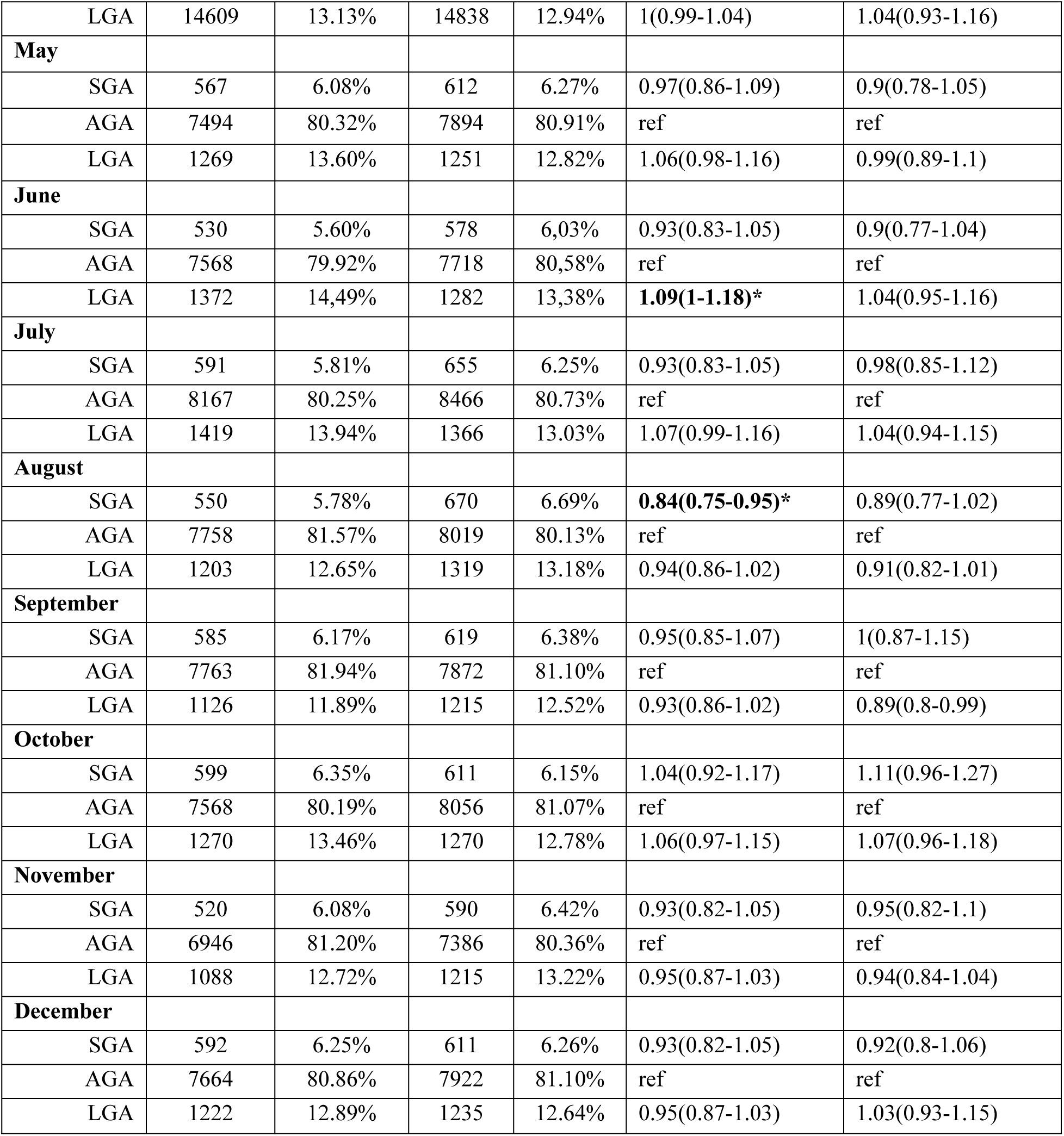
Proportion of birth weight newborns, Odds Ratios (CI95%) and adjusted Odd Ratio (CI95%) stratified by month of delivery in 2020 and 2019. Reference 2019 * p<0.05 ° *Adjustment for maternal nationality of origin, household incomes, single status, maternal education, maternal age, parity, maternal BMI and hypertension*.

## Discussion

We observed that, among mothers who gave birth in 2020 compared with 2019, there was an overall increase in the prevalence of HIP. No statistically significant differences were observed among women delivering between January and June, whereas a significant increase became apparent among those delivering from July onwards.

Importantly, the temporal pattern of HIP prevalence appeared to mirror the evolution of pandemic-related restrictions in Belgium. Women delivering in July were most likely screened for gestational diabetes in April, corresponding to the period of highest SI values during the first national lockdown. When monthly HIP adjusted ORs were reassigned to the corresponding month of screening, we observed a strong positive correlation between HIP prevalence and the SI. To our knowledge, this is the first study to directly examine the relationship between the intensity of governmental containment measures and temporal variations in HIP prevalence during the COVID-19 pandemic.

Previous studies have reported increases in gestational diabetes prevalence during the COVID-19 pandemic (1–4). Consistent with these findings, we observed a statistically significant increase in HIP prevalence, with effect sizes comparable to those reported in prior studies, but while earlier studies generally compared pre-pandemic and pandemic periods, they were unable to evaluate whether fluctuations in disease occurrence corresponded to changes in the intensity of containment measures. In contrast, our findings suggest that there may be a close temporal association between restriction intensity and HIP prevalence.

Several mechanisms could explain this association. Periods of stricter restrictions were characterized by reduced opportunities for physical activity, disruptions in daily routines, changes in dietary habits, and increased social isolation. These factors have all been linked to impaired glucose metabolism and increased gestational diabetes risk (2,4,8). In addition, psychological stress and anxiety were particularly elevated during lockdown periods and may have contributed through neuroendocrine pathways involving cortisol secretion and increased insulin resistance (1,3,4,25).

Conversely, some countries implemented alternative gestational diabetes screening strategies during the pandemic to reduce contact between pregnant women and healthcare facilities. These alternative strategies generally had lower sensitivity than standard oral glucose tolerance test-based screening and were associated with lower reported GDM prevalence (23). Similar modifications may have been implemented in some Belgian centres, although systematic information on their use is unavailable. If present, however, such changes would be expected to bias HIP ascertainment towards underdiagnosis rather than overdiagnosis. Consequently, the true increase in HIP prevalence may have been underestimated.

Interestingly, the association between the pandemic period and HIP persisted after adjustment for maternal BMI. Although the proportion of women classified as obese increased slightly in 2020, the shift in BMI distribution was modest and unlikely to fully explain the observed increase in HIP prevalence.

Given the established relationship between maternal hyperglycaemia and excessive fetal growth, we expected that an increase in HIP prevalence, coupled with restricted access to healthcare services, might result in a higher prevalence of large-for-gestational-age (LGA) newborns. This was not observed. The distribution of birthweight percentiles remained remarkably stable between 2019 and 2020, and the only statistically significant finding was a small reduction in LGA prevalence in September 2020, which may represent a chance finding.

The absence of an increase in LGA newborns despite a higher prevalence of gestational diabetes has also been reported by other authors (16,25,26). One possible explanation is that the rapid adoption of telemedicine improved diabetes follow-up and glycaemic control during pregnancy, therefor the impact of GDM on fetal growth was hypothesised to be mitigated by intensive management and earlier diagnosis (3,25).

Another possibility is that the increase in HIP prevalence was primarily driven by an increase in mild forms of gestational diabetes. Such cases may have been sufficient to increase the prevalence of diagnosed HIP without generating a measurable effect on fetal growth and birthweight distribution at the population level. Unfortunately this hypothesis could not be tested in our study since we relied on birth registry data without access to individual patients’ glycaemic measurements.

### Strengths and limitations

The strengths of this study include its nationwide population-based design, encompassing all singleton births in Belgium over a two-year period, and the availability of systematically collected sociodemographic and clinical covariates. The use of a comprehensive birth registry minimized selection bias and allowed adjustment for a broad range of potential confounders.

Several limitations should nevertheless be acknowledged. First, the observational nature of the study precludes causal inference. Although a strong temporal correlation was observed between HIP prevalence and SI, the study design cannot establish that containment measures directly caused the increase in HIP. Second, HIP status was derived from administrative data and did not allow differentiation between gestational and pregestational diabetes, although the latter remains relatively uncommon in this population. Third, because HIP was recorded at delivery, the timing of gestational diabetes screening had to be approximated according to contemporaneous clinical guidelines. Fourth, residual confounding cannot be excluded, particularly with respect to factors not available in the registry.

## Conclusions

During the COVID-19 pandemic, we observed a progressive increase in the prevalence of hyperglycaemia in pregnancy in Belgium. The temporal evolution of HIP prevalence seems to be strongly associated to the SI, with higher odds of HIP diagnosis observed during periods of more restrictive containment measures. These findings suggest that societal restrictions may have had measurable indirect effects on metabolic health during pregnancy.

Despite the increase in HIP prevalence, no corresponding increase in large- or small-for-gestational-age newborns was observed. Future studies should investigate the mechanisms linking containment measures and maternal glucose metabolism, as well as the potential impact of these changes on other maternal and neonatal outcomes.

More broadly, understanding how maternal health conditions respond to periods of social disruption and restricted mobility may help anticipate unintended consequences of future public health emergencies requiring similar containment measures.

## Data Availability

The data that support the findings of this study were obtained from the Belgian Statistical Office (Statbel) and are subject to legal and ethical restrictions. The data are not publicly available. Access to the data may be requested from Statbel and the Institutional Review Board of the Belgian Statistical Office, subject to approval and compliance with Belgian data protection regulations. Aggregated data and statistical code are available from the corresponding author upon reasonable request.

## Acknowledgements

We thank the Belgian Statistical Office (Statbel) for providing the data and Dr Ariana Keramyda for the helpful language review.

## Author Contributions

EC designed the study, analyzed the data, and wrote the main manuscript. AV designed the study and reviewed the manuscript, SA contributed to the study design, results interpretation and MB contributed to the study design, results interpretation, figures design and reviewed the manuscript, SD designed the study and reviewed the manuscript, CL reviewed the manuscript, AD contributed to the study design, results interpretation and reviewed the manuscript, JR designed the study, retrieved data, analised the data and reviewed the manuscript

## Abbreviations

aOR: Adjusted Odds Ratio
AUDIPOG: Association des Utilisateurs de Dossiers Informatisés en Pédiatrie, Obstétrique et Gynécologie
BMI: Body Mass Index
CEpIP: Centre d’Épidémiologie Périnatale (Center for Perinatal Epidemiology)
CI: Confidence Interval
COVID-19: Coronavirus Disease 2019 GD Gestational Diabetes
HIP: Hyperglycemia in Pregnancy
HT: Hypertension
IADPSG: International Association of Diabetes and Pregnancy Study Groups
LGA: Large for Gestational Age
OR: Odds Ratio
SGA: Small for Gestational Age
SI: Stringency Index
SPE: Studiecentrum voor Perinatale Epidemiologie (Study Centre for Perinatal Epidemiology)

## Notes

### Competing Interest Statement

The authors have declared no competing interest.

### Funding Statement

The author(s) received no specific funding for this work.

### Author Declarations

This study is based on anonymous administrative data and is expempt from going through ethics commettee. This study and the use of related data were approved by the Institutional Review Board (IRB) of the Belgian Statistical Office (Statbel): reference number 2022/052. (https://statbel.fgov.be/sites/default/files/files/documents/Over%20Statbel/Microdata_EN.pdf). Participant consent is the responsibility of Statbel in accordance with Belgian legislation. (https://statbel.fgov.be/en/about-statbel/privacy/privacy-gdpr).

